# Serum Vitamin D Status and Risk of Early and Late Dental Implant Failure: A Systematic Review of Multivariate-Adjusted Primary Studies with Explicit Vitamin D Cut-off Classification

**DOI:** 10.64898/2026.01.05.26343458

**Authors:** Beatriz Pardal-Peláez, José Luis Pardal-Refoyo

**Affiliations:** Department of Surgery, Dentistry area, Faculty of Medicine and Dental Clinic, University of Salamanca, Salamanca, Spain; Department of Surgery, Otorhinolaryngology Area, Otorhinolaryngology and Head and Neck Surgery, University Hospital of Salamanca, Faculty of Medicine, University of Salamanca, Salamanca, Spain

## Abstract

**Statement of problem:** Dental implant failure remains a significant clinical concern, with early loss often attributed to impaired osseointegration. Recent research has considered the role of serum vitamin D in implant integration, yet the precise relationship between vitamin D status and implant loss, particularly when distinguishing early and late failures, is not fully established.

**Purpose:** The objective of this analysis was to evaluate the association between explicit serum vitamin D cut-off values and clinically confirmed dental implant failure, with a particular focus on differentiating early (pre-loading) from late (post-loading) failures. The review also sought to determine whether primary studies used multivariate adjustment for potential confounders.

**Material and methods:** The draft of this revision was registered in PROSPERO (CRD420251049631, https://www.crd.york.ac.uk/PROSPERO/view/CRD420251049631). A comprehensive literature search was conducted using AI-assisted tools to identify primary research studies, including randomized controlled trials and cohort studies, published in English, Spanish, French, German, or Italian. Eligible studies required explicit vitamin D threshold categorization, clinically verified implant loss, and clear differentiation of early and late failures. Data extraction included study design, vitamin D categorization, analytical methods, and outcomes.

**Results:** Identified studies predominantly consisted of retrospective and prospective cohorts examining early implant failures, frequently using cut-offs such as >30 ng/mL, 10–30 ng/mL, and <10 ng/mL for serum vitamin D. Results suggested a higher frequency of early failures in individuals with severe vitamin D deficiency; however, all studies relied on univariate analyses without multivariate adjustment for confounders. Late implant failures were rarely addressed.

**Conclusions:** Current evidence indicates a possible association between low serum vitamin D and early dental implant failure, but the lack of robust statistical adjustment prevents definitive conclusions. High-quality studies with rigorous confounder control and explicit early versus late failure analysis are needed.

## INTRODUCTION

Early loss of dental implants remains a significant clinical challenge. Current research distinguishes between early (pre-load) and late (post-load) failures, and some studies have controlled for confounding factors, such as smoking and periodontal disease, through multivariate adjustments.^1,2^

Insufficient vitamin D can negatively impact osseointegration and increase the risk of early dental implant loss, though studies on this issue show mixed results and have methodological limitations.^1–3^

25-hydroxyvitamin D is the biomarker used to assess vitamin D status, with clinical cut-off points indicating deficiency when it is less than 20 ng/mL, insufficiency between 20 and 30 ng/mL, and sufficiency when it is greater than 30 ng/mL.^2,4^

Understanding the association between low vitamin D levels and dental implant failure is crucial for risk stratification and optimization of outcomes in dental implantology. Preoperative Vitamin D evaluation and supplementation may improve clinical outcomes in patients with subphysiological vitamin D levels.^2,4^

Research Gap. Despite widespread recognition of vitamin D deficiency among oral surgery patients, there remains limited evidence clarifying its direct impact on dental implant survival. Current literature primarily focuses on describing the prevalence of vitamin D deficiency or exploring associations with biological markers related to peri-implant health, rather than providing definitive data on how specific serum 25(OH)D thresholds influence the rates of clinically confirmed early or late implant loss ^5^. Furthermore, there is emerging but underexplored concern about the potential adverse effects of excessive vitamin D (hypervitaminosis D) on implant outcomes.^6^ Objective biomechanical assessments, such as resonance frequency analysis in vitamin D-deficient patients, are also relatively new and have not been systematically integrated into outcome studies.^7^

Rationale. Given the essential role of vitamin D in bone metabolism and the high prevalence of deficiency—and, in rare cases, excess—among dental implant candidates, defining the exact relationship between vitamin D status and implant failure is crucial for improving risk stratification and patient management. A clearer understanding could enable tailored interventions, both in prevention and during the peri-implant period, for those at greatest risk of implant complications.^5,6^

Objectives. The objective of this research is to evaluate the association between explicit serum 25(OH)D cut-off values and the incidence of clinically confirmed dental implant failure, including both early (pre-loading) and late (post-loading) events. The review also sought to determine whether primary studies used multivariate adjustment for potential confounders.

## MATERIAL AND METHODS

The draft of this revision was registered in PROSPERO (CRD420251049631, https://www.crd.york.ac.uk/PROSPERO/view/CRD420251049631)^8^.

This scoping review was conducted and reported in accordance with PRISMA guidelines (http://www.prisma-statement.org). The research aimed to systematically map available primary literature on the association between serum vitamin D cut-off values and dental implant failure, both early (pre-loading) and late (post-loading). A comprehensive search strategy was developed utilizing multiple international scientific databases, including PubMed (https://pubmed.ncbi.nlm.nih.gov), Embase (https://www.embase.com), Web of Science (https://www.webofscience.com), the Cochrane Central Register of Controlled Trials (https://www.cochranelibrary.com/central), and OpenGrey (http://www.opengrey.eu). Searches were conducted in English, Spanish, French, German, and Italian.

The search strategy combined the following terms and Boolean operators: (“vitamin D” OR “25-hydroxyvitamin D” OR “cholecalciferol”) AND (“dental implant” OR “oral implant”) AND (“failure” OR “loss” OR “survival”) AND (“early” OR “late”) AND (“cut-off” OR “threshold” OR “deficiency”) AND (“multivariate” OR “confounder” OR “adjusted”). MeSH terms and database-specific subject headings were also used where applicable.

Both researchers participated in the identification and selection of articles.

Inclusion criteria encompassed primary clinical studies (randomized controlled trials, prospective or retrospective cohorts, and case–control studies) that explicitly defined and compared serum 25-hydroxyvitamin D cut-off values, reported clinically verified dental implant loss, distinguished between early and late failures based on author definitions, employed multivariate analysis for confounders, and were published in the specified languages. Exclusion criteria were non-primary studies (reviews, animal/in vitro work), studies lacking explicit vitamin D cut-offs, absence of early/late failure distinction, missing confounder adjustment, or publication in other languages.

Data extraction was performed using an AI-assisted platform, Undermind (https://undermind.ai), in both the selection and screening phases and for automated data extraction and preliminary synthesis. Candidate studies were screened for eligibility and relevant data were captured on study design, sample size, cut-off values for vitamin D, analytic methods, outcomes, and adjustment for confounders.

The primary variables analyzed were serum 25-hydroxyvitamin D concentration (measured in ng/mL, categorized using study-specific thresholds), implant failure (early and/or late, as defined by each study), and covariates related to patient or treatment factors. Owing to the predominance of descriptive data, the principal statistical approach was qualitative synthesis. Where available, reported statistical tests (such as chi-square tests or logistic regression) from individual studies were noted.

In methodologically homogeneous studies, meta-analyses were performed on the results of dental implant failures related to vitamin D levels. The statistical program Jamovi version 2.6.17.0 was used (The jamovi project (2024). *jamovi*. (Version 2.6) [Computer Software]. Retrieved from https://www.jamovi.org; R Core Team (2024). *R: A Language and environment for statistical computing*. (Version 4.4) [Computer software]. Retrieved from https://cran.r-project.org. (R packages retrieved from CRAN snapshot 2024-08-07).

## RESULTS

A total of 10 articles were identified through the comprehensive search strategy^5– 7,9–15^. After title and abstract screening, 4 records were excluded for not meeting the inclusion criteria. Full-text assessment led to the exclusion of 2 additional articles: one due to the absence of explicit vitamin D cut-off reporting, and another owing to lack of failure timing distinction. Ultimately, 4 studies met all eligibility criteria for qualitative synthesis^9–12^. The PRISMA flowchart outlining this process is shown in Figure 1.

**Figure 1.**
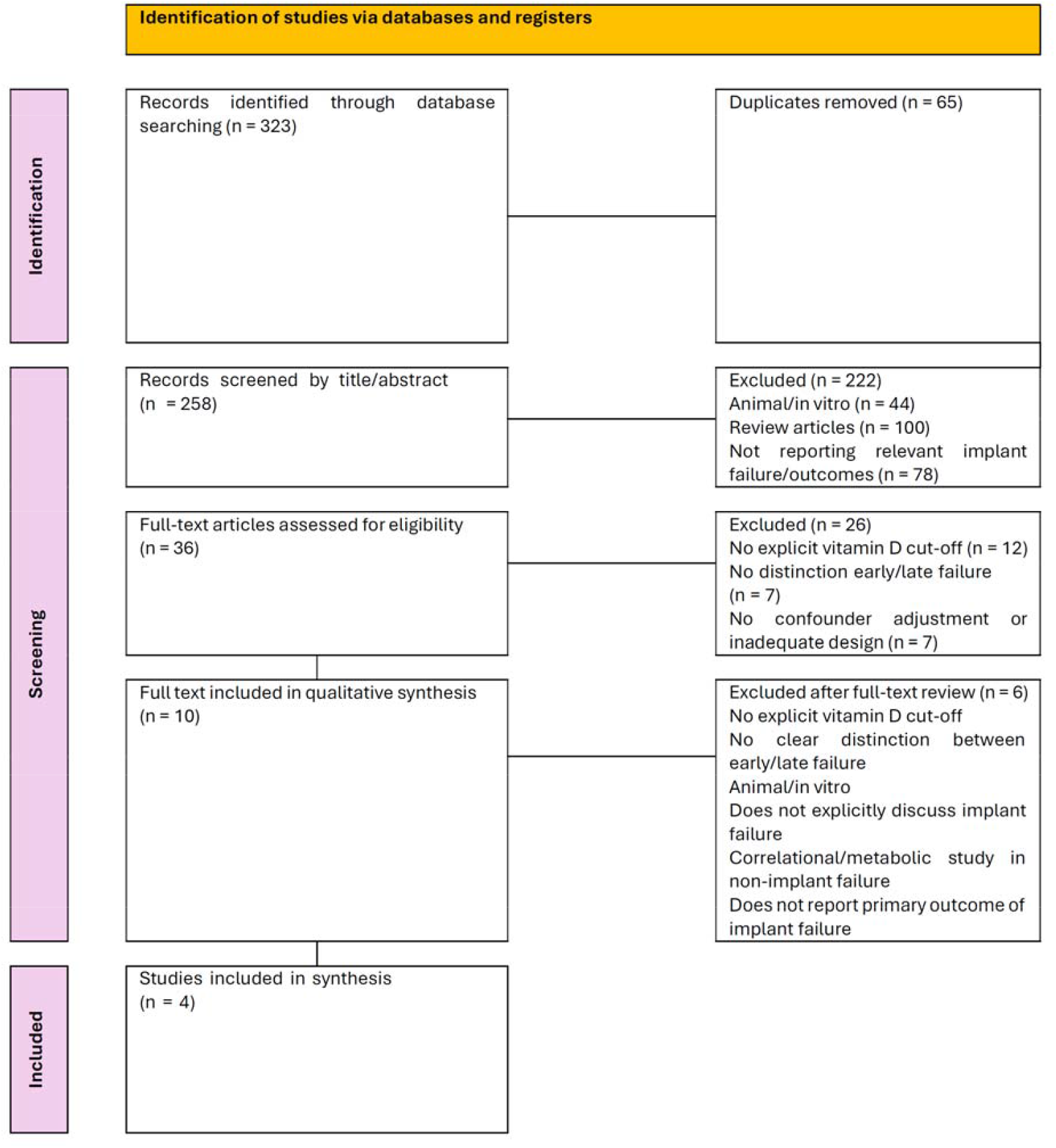
PRISMA 2020 flow diagram (Source: Page MJ, et al. BMJ 2021;372:n71. doi: 10.1136/bmj.n71).

Table 1 (see Annex) shows the data obtained from the studies analyzed.

**Table 1.** Characteristics and Main Results of Included Studies.

| Reference | Method | N patients | N implants | Hypovitaminosis | Deficiency | Insufficient | Sufficient | Failures in Hypovitaminosis | Failures in Patients with Normal Vitamin D | Early Failures (<10 ng/mL Vitamin D) | Early Failures (10-30 ng/mL Vitamin D) | Early Failures (>30 ng/mL Vitamin D) | Total failures |
| --- | --- | --- | --- | --- | --- | --- | --- | --- | --- | --- | --- | --- | --- |
| Bawa et al., 2022 | P | 100 | 100 | 77 | 6 | 71 | 23 | 6 | 1 | 1 | 5 | 1 | 7 |
| Mangano et al., 2016 | R | 822 | 1625 | 428 | 22 | 406 | 394 | 18 | 9 | 2 | 16 | 9 | 27 |
| Mangano et al., 2018 | R | 885 | 1740 | 475 | 27 | 448 | 410 | 23 | 12 | 3 | 20 | 12 | 35 |
| Mohsen et al., 2024 | P | 53 | 143 | 42 | 6 | 36 | 11 | 8 | 2 | 6 | 2 | 2 | 10 |

Hypovitaminosis D (<30 ng/mL); Insufficient Vitamin D (10-30 ng/mL); P: prospective; R: retrospective; Sufficient Vitamin D (>30 ng/mL); Vitamin D Deficiency (<10 ng/mL)

All included studies focused on early implant failure, defined as loss occurring before prosthetic loading, typically within 3–4 months following placement. Vitamin D was categorized into three major strata: >30 ng/mL (sufficient), 10–30 ng/mL (insufficient), and <10 ng/mL (deficient). Across studies, a trend toward a higher frequency of early failures in severely deficient vitamin D groups was observed in retrospective cohorts, whereas prospective studies yielded inconsistent results.

All studies reported results based on the number of implant failures relative to the number of patients. Only the study by Mohsen et al., 2024 ^12^ stratified the results of implant number failures according to vitamin D levels (in the Deficient group (<10 ng/mL) 6/13 = 0.46 [IC 95% 0.23-0.71]; in the Insufficient group (10-30 ng/mL) 2/86 = 0.023 [IC 95% 0.0019-0.087] and in the Sufficient group (>30 ng/mL) 2/42 = 0.048 [IC 95% 0.0055-0.17].

The magnitude of observed effect varied; early failure rates rose from 0.0286 [IC 95% 0.0192-0.0426] in the sufficient group to 0.20 [IC 95% 0.12-0.32] in the severely deficient group as shown in Table 2 (see Annex). However, none of the studies applied multivariate statistical adjustment for confounders such as age, smoking status, systemic conditions, or surgical factors. As a result, only univariate associations were reported, commonly using chi-square tests without reporting p-values or confidence intervals.

**Table 2.** Implant failure in the groups analyzed.

| Vit. D level (ng/mL) Group | N Patients | N implant failures | Estimate | CI Lower Bound | CI Upper Bound | P<sub>adjusted</sub> | SE<sub>proportion</sub> |
| --- | --- | --- | --- | --- | --- | --- | --- |
| Patients with low vitamin D (<30 ng/mL) | 1022 | 55 | 0.0486 | 0.035 | 0.062 |  |  |
| (<10 ng/mL Vitamin D) | 61 | 12 | 0.1967 | 0.1154 | 0.3153 | 0.4706 | 0.1211 |
| (10-30 ng/mL Vitamin D) | 961 | 43 | 0.0447 | 0.0333 | 0.0599 | 0.0444 | 0.0217 |
| Patients with normal vitamin D (>30 ng/ml) | 838 | 24 | 0.0286 | 0.0192 | 0.0426 | 0.0870 | 0.0415 |
| Global | 1860 | 79 | 0.0381 | 0.029 | 0.047 |  |  |

Note. Tau<sup>2</sup> Estimator: Restricted Maximum-Likelihood Random-Effects Model (k = 4)

The dichotomous meta-analysis of dental implant failures in the selected studies in patients in the group with normal versus low vitamin D levels offers an absence of heterogeneity (I^2^ = 0 %; see Forest Plot in Figure 2) and low publication bias (see Funnel Plot in Figure 3).

**Figure 2.**
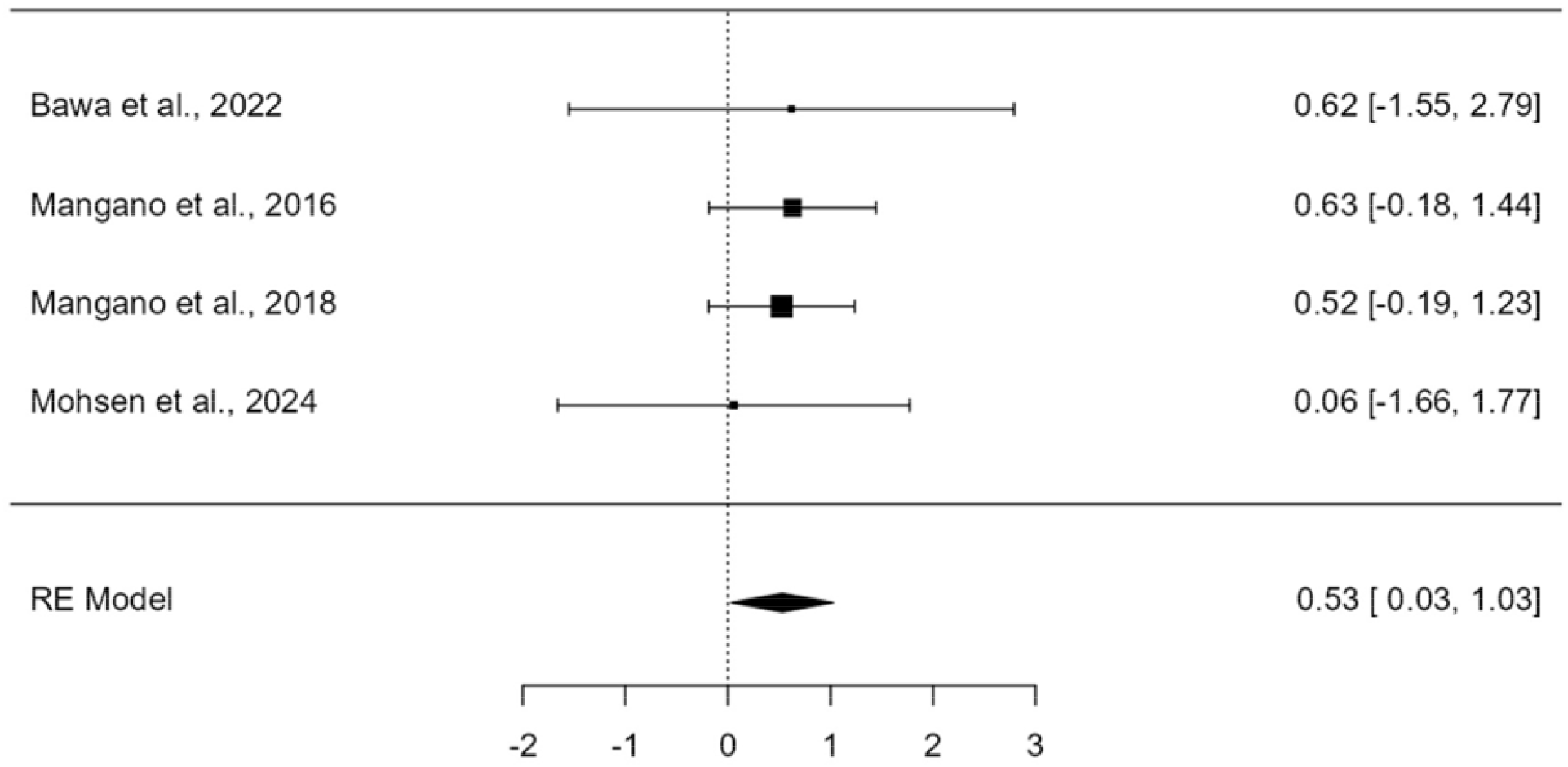
Forest Plot with the prevalence of the di□erence in dental implant failures in the groups with normal vitamin D (>30 ng/mL) versus the groups of patients with low vitamin D (<30 ng/mL).

**Figure 3.**
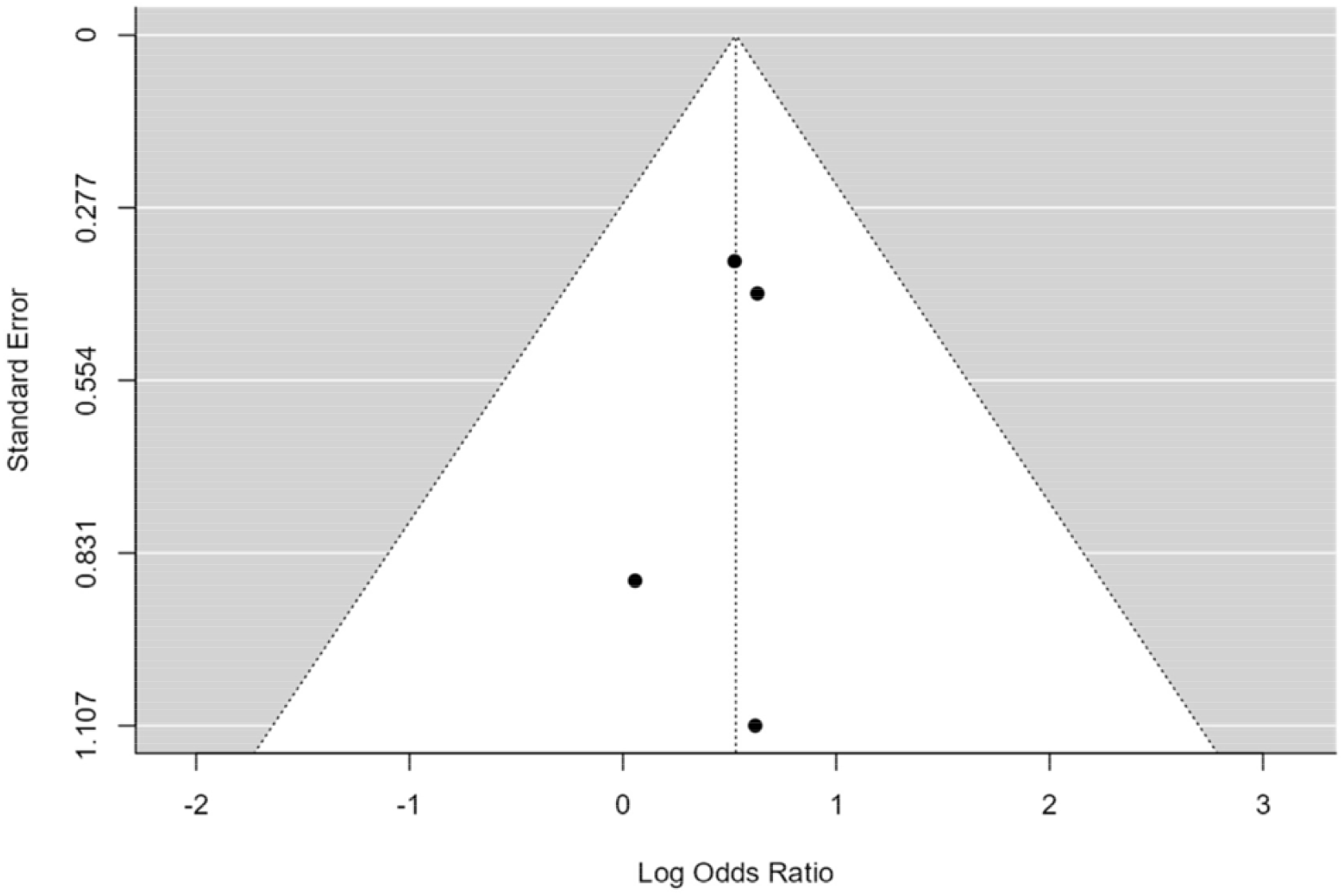
Funnel Plot showing low risk of publication bias.

Quality assessment using the Newcastle–Ottawa Scale for cohort studies indicated moderate risk of bias across all included studies (Table 3, see Annex). The main sources of bias were lack of control for confounding, incomplete reporting of patient selection, and short follow-up periods limited to early failures. According to the GRADE system (https://www.gradepro.org/), the overall quality of evidence was rated as low, with a weak strength of recommendation due to study design limitations and unexplained heterogeneity in results.

**Table 3.** Risk of Bias Assessment (Newcastle–Ottawa Scale) for Included Articles.

| Reference | Selection | Comparability | Outcome Assessment | Follow-up | Overall Risk of Bias |
| --- | --- | --- | --- | --- | --- |
| Mangano 2016 et al. | Moderate | Low | Moderate | Low | Moderate |
| Mangano 2018 et al. | Moderate | Low | Moderate | Low | Moderate |
| Bawa 2022 et al. | Moderate | Low | Moderate | Low | Moderate |
| Mohsen 2024 et al. | Moderate | Low | Moderate | Low | Moderate |

## DISCUSSION

The present review systematically evaluated primary research studies assessing the association between specific serum vitamin D cut-off values and clinically confirmed dental implant failure, with particular attention to the timing of failure and statistical adjustment for confounding variables. The overall findings indicate a possible association between severe vitamin D deficiency and an increased rate of early dental implant failure, as higher failure rates were observed in patients with vitamin D levels below 10 ng/mL compared to those with sufficient levels in large retrospective cohorts.^9,10^ However, this trend was not consistent across smaller prospective cohorts, where failures also occurred in patients with sufficient vitamin D or did not exhibit a clear relationship with deficiency.^11,12^

When compared with previous research on vitamin D and bone metabolism, these results align with the biological premise that vitamin D plays a role in osseointegration and bone remodeling, potentially impacting initial implant stability and integration.^10^ Nevertheless, the inability of any identified study to perform multivariate adjustment for major confounders such as age, systemic diseases, smoking, or surgical factors significantly limits the interpretability and causal inference of the findings. This absence of robust statistical control is a notable gap, as unmeasured confounding may explain the observed associations.^9–12^

Clinically, although low vitamin D levels are suspected to increase early implant loss risk, the evidence base is not sufficiently rigorous to guide routine vitamin D screening or supplementation solely for implant success without further high-quality research. The findings do not address late dental implant failures or outcomes, leaving a significant gap regarding the role of vitamin D beyond osseointegration. Moreover, methodological heterogeneity—differences in vitamin D cut-offs, assay techniques, study populations, and outcome definitions—further restricts generalizability.

Strengths of this review include the use of explicit selection criteria, inclusion of studies employing recognized vitamin D thresholds, and focus on clinically confirmed implant loss. Opportunities for further research include well-designed prospective cohort studies or randomized controlled trials that incorporate multivariate analysis to adjust for confounders, standardized outcome definitions distinguishing early and late failures, and extended follow-up. Larger sample sizes and direct comparison of supplementation interventions may also clarify the dose–response relationship and clinical utility of vitamin D in dental implantology.^9,10^

Several key confounding factors should be evaluated when considering outcomes in implantology identified in the reviewed literature. Patient-related factors include age, sex, body mass index (BMI), smoking habits, presence of diabetes mellitus, systemic bone conditions, medication use, and history of periodontitis. Implant-related factors encompass the location of the implant, dimensions, treatment surface and process, whether immediate or delayed loading is applied, and the follow-up period. Surgical factors involve whether previous surgeries such as bone grafting or sinus lifts have been performed, the quality and quantity of the recipient bone, the surgeon’s experience, and the surgical protocol—including drilling technique, type of flap, and anaesthesia used. Other important considerations are the station and year of vitamin D measurement, the type of assay used to measure vitamin D, and periodontic characteristics such as the type of rehabilitation and type of load applied.

Limitations of this review reflect those of the included studies: observational designs, short follow-up periods focused on early failures, lack of blinding, and the absence of adjustment for relevant covariates. The limited number of eligible studies and their methodological deficiencies result in a low level of evidence and weak recommendation strength according to GRADE criteria.

Future research should aim to fill these gaps by implementing robust causal models, longitudinal designs, and standardized variables to provide more definitive guidance for clinical practice.

## CONCLUSIONS

1. There is a potential association between severe vitamin D deficiency and an increased risk of early dental implant failure, but this relationship is not consistent across all study designs.
2. Existing studies focus predominantly on early failures and lack robust multivariate adjustment for confounding factors, limiting the ability to draw definitive causal conclusions.
3. The evidence base does not support changes in clinical practice regarding vitamin D screening or supplementation solely for dental implant success at this time.
4. Significant methodological limitations, including small sample sizes, observational designs, and absence of standardized outcome measures, reduce the overall quality of available evidence.
5. Further high-quality prospective studies with standardized definitions, rigorous confounder adjustment, and extended follow-up are needed to clarify the role of vitamin D in dental implant outcomes.

## Data Availability

All data produced in the present work are contained in the manuscript

https://www.crd.york.ac.uk/PROSPERO/view/CRD420251049631

## ANNEX

## REFERENCES

1. Buzatu BLR, Buzatu R, Luca MM. Impact of Vitamin D on Osseointegration in Dental Implants: A Systematic Review of Human Studies. Nutrients. 2024;16:209.

2. Wiedemann TG, Jin HW, Gallagher B, Witek L, Miron RJ, Talib HS. Vitamin D Screening and Supplementation-A Novel Approach to Higher Success: An Update and Review of the Current Literature. J Biomed Mater Res B Appl Biomater. 2025;113:e35558.

3. Bazal-Bonelli S, Sánchez-Labrador L, Cortés-Bretón Brinkmann J, Cobo-Vázquez C, Martínez-Rodríguez N, Beca-Campoy T, et al. Influence of Serum Vitamin D Levels on Survival Rate and Marginal Bone Loss in Dental Implants: A Systematic Review. Int J Environ Res Public Health. 2022;19:10120.

4. Al-Quisi AF, A Jamil F, M Al-Anee A, Jassim Muhsen S. Relationship Between the Level of Vitamin D3 Deficiency and Successful Osseointegration: A Prospective Clinical Study. Sci World J. 2024;2024:9933646.

5. Waskiewicz K, Oth O, Kochan N, Evrard L. Des facteurs de risque généralement négligés en chirurgie orale et en implantologie :le taux élevé de LDL-cholestérol et le taux insuffisant de la vitamine D. Rev Med Brux. 2018;39:70–7.

6. Cheng YC, Murcko L, Benalcazar-Jalkh EB, Bonfante EA. Hypervitaminosis D is correlated with adverse dental implant outcomes: A retrospective case-control study. J Dent. 2024 Aug;147:105137.

7. Toy V, Sabancı A. Resonance frequency analysis of dental implants in patients with vitamin D deficiency. Clin Oral Investig. 2024;28:682.

8. Pardal-Refoyo JL, Pardal-Pelaéz B. Serum Vitamin D Status and Risk of Early and Late Dental Implant Failure: A Systematic Review of Multivariate-Adjusted Primary Studies with Explicit Vitamin D Cut-off Classification. PROSPERO; 2025. Available from: https://www.crd.york.ac.uk/PROSPERO/view/CRD420251049631

9. Mangano F, Mortellaro C, Mangano N, Mangano C. Is Low Serum Vitamin D Associated with Early Dental Implant Failure? A Retrospective Evaluation on 1625 Implants Placed in 822 Patients. Mediators Inflamm. 2016;2016:5319718.

10. Mangano FG, Oskouei SG, Paz A, Mangano N, Mangano C. Low serum vitamin D and early dental implant failure: Is there a connection? A retrospective clinical study on 1740 implants placed in 885 patients. J Dent Res Dent Clin Dent Prospects. 2018;12:174–82.

11. Bawa S, Sharma P, Jindal V, Malhotra R, Malhotra D, Chauhan P. A clinico-relationship between Vitamin D and early implant failure. IP Int J Periodontol Implantol. 2022;7:15–22.

12. Mohsen K, AbdEl-Raouf MN, Makram K, Elkassaby M, Khairy MA, Mahmoud AbdelAziz, et al. Is Vitamin D Deficiency a Risk Factor for Osseointegration of Dental Implants - A Prospective Study. Ann Maxillofac Surg. 2024;14:21–6.

13. Lin G, Ye S, Liu F, He F. A retrospective study of 30,959 implants: Risk factors associated with early and late implant loss. J Clin Periodontol. 2018;45:733–43.

14. Savchuk O, Krasnov V, Yurzhenko A, Far SA. Predicting the success of dental implantation in patients with defects in the dentition against the background of chronic generalized periodontitis. Actual Dent. 2021;5:64–6.

15. Francis J, Barber HD, Beals D, Siu T. The Relationship of low-serum Vitamin D and Early Dental Implant Failure. J Oral Implantol. 2024;50:215–8.

